# Coffee consumption is associated with later age-at-onset of Parkinson’s disease

**DOI:** 10.1101/2025.02.07.25321819

**Authors:** Dariia Kuzovenkova, Lang Liu, Ziv Gan-Or, Konstantin Senkevich

**Author notes:** **Corresponding author:** Konstantin Senkevich, Department of Neurology and Neurosurgery, McGill University, 1033 Pine Avenue, West, Ludmer Pavilion, room 309, Montreal, QC, H3A 1A1, Canada.

## Abstract

Observation studies suggest that coffee consumption may lower the risk of Parkinson’s disease (PD). The aim of this study was to explore the causal relationship and genetic association between coffee consumption and the age-at-onset (AAO), risk, and progression of PD. Using Mendelian randomization, we identified a significant association between coffee consumption and delayed PD AAO (IVW: OR, 1.91; 95% CI 1.53–2.38; P=8.072e-09), but no causal association or genetic correlation with PD risk or progression. Our findings suggest a potential protective causal effect of higher coffee consumption on PD AAO, with no evidence of an association with PD risk or progression.

## Introduction

Prospective studies over the past two decades have consistently associated caffeine consumption with a reduced risk of Parkinson’s disease (PD) ^1^. Additionally, lower levels of caffeine and its metabolites have been observed in PD patients compared to controls ^2^. Although a clinical trial has not demonstrated significant symptomatic benefits of caffeine intake for PD ^3^, its potential to delay motor symptom onset may create an apparent protective effect.

The potential protective effects of caffeine in PD may involve multiple mechanisms, including antagonism of adenosine receptors and dopaminergic modulation ^4^. Observational studies further support protective effects demonstrating a later age at onset (AAO) of PD in analyses of coffee and tea consumption ^5, 6^. Despite previous Mendelian randomization (MR) studies failed to identify a causal link between coffee consumption and PD risk ^7, 8^, effect on AAO and clinical progression remains underexplored. In addition, no strong genetic link between caffeine consumption and sporadic PD that would explain protective association has been demonstrated to date.

To address the gaps in understanding the relationship between caffeine consumption and PD, we aimed to investigate the genetic and causal associations between the amount of coffee intake and PD AAO, risk, and progression. Using MR and genetic correlation, we assessed the relationship between coffee consumption and PD outcomes. We further constructed polygenic risk score (PRS) for coffee consumption and studied association with PD risk and AAO.

## Methods

### Study population

We used publicly available genome-wide association study (GWAS) data to explore the genetic and causal relationships between caffeine consumption and PD AAO, risk, and progression. Independent GWAS significant single nucleotide polymorphisms (SNPs) associated with caffeine consumption were obtained from the UK Biobank GWAS data (Data field ID 1498) ^9^ **(Table 1)**. As an outcome, we used the largest available GWAS statistics on PD risk with 27,693 participants of European ancestry (PD cases = 15,056; controls = 12,637) ^10^. UK Biobank data was excluded from it to prevent overlapping samples that could introduce bias. We also used GWAS results for PD AAO in the form of a continuous variable with 17,996 PD patients of European ancestry ^11^. To examine coffee consumption’s effects on PD symptom progression (UPDRS Part III), cognitive decline (MoCA, MMSE), insomnia, and hyposmia, we used publicly available GWAS summary statistics data for these traits ^12^. For the genetic correlation analysis, we used a caffeine consumption GWAS for which complete summary statistics was available ^13^.

**Table 1.** GWAS summary statistics utilized in the MR analysis.

| Studied trait | Study | Sample size, N | Type of trait |
| --- | --- | --- | --- |
| Mendelian Randomization exposure |  |  |  |
| Coffee consumption | PMID: 31412118 <sup>9</sup> | 408,191 | Continuous |
| Mendelian Randomization outcome |  |  |  |
| PD risk | PMID: 31701892 <sup>10</sup> | Cases = 15,056;<br>controls = 12,637 | Binary |
| PD AAO | PMID: 30957308 <sup>11</sup> | 17,996 | Continuous |
| UPDRS3 | PMID: 31505070 <sup>12</sup> | 1,398 | Continuous |
| MMSE |  | 1,329 | Continuous |
| MoCA |  | 1,000 | Continuous |
| Hyposmia |  | 1,027 | Continuous |
| Sleep |  | 1,136 | Continuous |
| Genetic correlation |  |  |  |
| Coffee consumption | PMID: 33287642 <sup>13</sup> | 362,316 | Continuous |

PD - Parkinson disease; AAO - Age at Onset; UPDRS3 - unified Parkinson's disease rating scale part 3; MMSE - Mini Mental State Examination; MoCA - Montreal Cognitive Assessment;

### Mendelian randomization and genetic correlation

MR analysis was used to assess the causal relationship between caffeine consumption and PD traits. Single nucleotide polymorphisms (SNPs) significantly associated with caffeine consumption (P < 5 × 10□ □) were selected as instrumental variables (IVs). SNPs were clumped using linkage disequilibrium thresholds (r^2^ < 0.001, 10,000 kb window) to ensure independence, and IV strength was evaluated using F-statistics (threshold ≥ 10). The inverse variance-weighted (IVW) method was the primary MR approach, with MR-Egger and weighted median estimates as sensitivity analyses ^14^. Directional pleiotropy was assessed using the MR-Egger intercept test, and heterogeneity was evaluated with Cochran’s Q test. Pleiotropic SNPs were identified and excluded using MR-PRESSO ^15^. Genetic correlation between caffeine consumption and PD traits was calculated using linkage disequilibrium score regression ^16^.

### Polygenic risk score

We evaluated the cumulative genetic contribution of caffeine consumption–associated SNPs to PD risk by constructing a PRS from genome-wide significant variants associated with caffeine intake ^9^. Linkage disequilibrium (LD) clumping (r^2^ < 0.1, 250 kb window) was applied, and scores were adjusted for age, sex, and principal components. The PRS was then tested across seven independent cohorts for both PD risk and AAO **(Supplementary Table 1)**, followed by a meta-analysis of the combined results.

## Results

We performed MR analysis of coffee consumption and PD AAO using 28 SNPs as IVs. The initial results showed no causal association (IVW: OR, 2.260; 95% CI, 0.568–8.989; P = 0.247). However, MR-PRESSO identified 16 pleiotropic SNPs **(Supplementary Table 2)**, which were excluded from a subsequent analysis. After removing pleiotropic SNPs, MR analysis with the remaining 12 SNPs revealed a strong causal association between coffee consumption and PD AAO (IVW: OR, 1.909; 95% CI, 1.532–2.377; P = 8.072e-09; **Figure 1; Table 2)**. Because AAO is a continuous outcome, OR > 1 indicates that greater coffee consumption is causally linked to a later onset of PD. Sensitivity analyses confirmed the robustness of these findings. No residual pleiotropy was detected (MR-PRESSO: P = 0.202; Egger: P = 0.851), and heterogeneity tests showed no evidence of inconsistency (Cochran’s Q for IVW: P = 0.074; **Supplementary Table 3**).

**Table 2.**
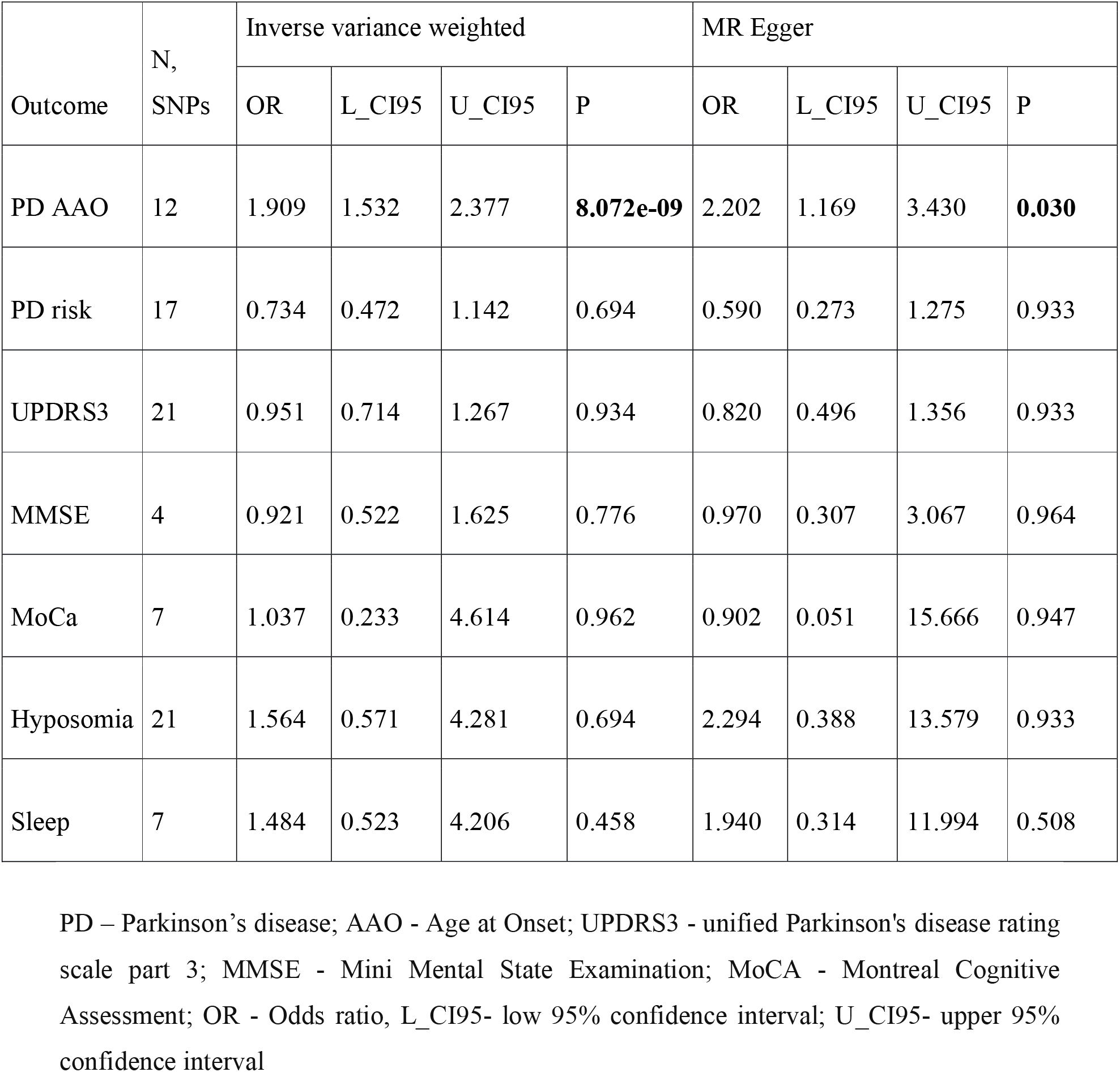
Mendelian randomization estimates of genetically predicted coffee consumption on Parkinson’s disease risk, AAO and progression.

**Figure 1.**
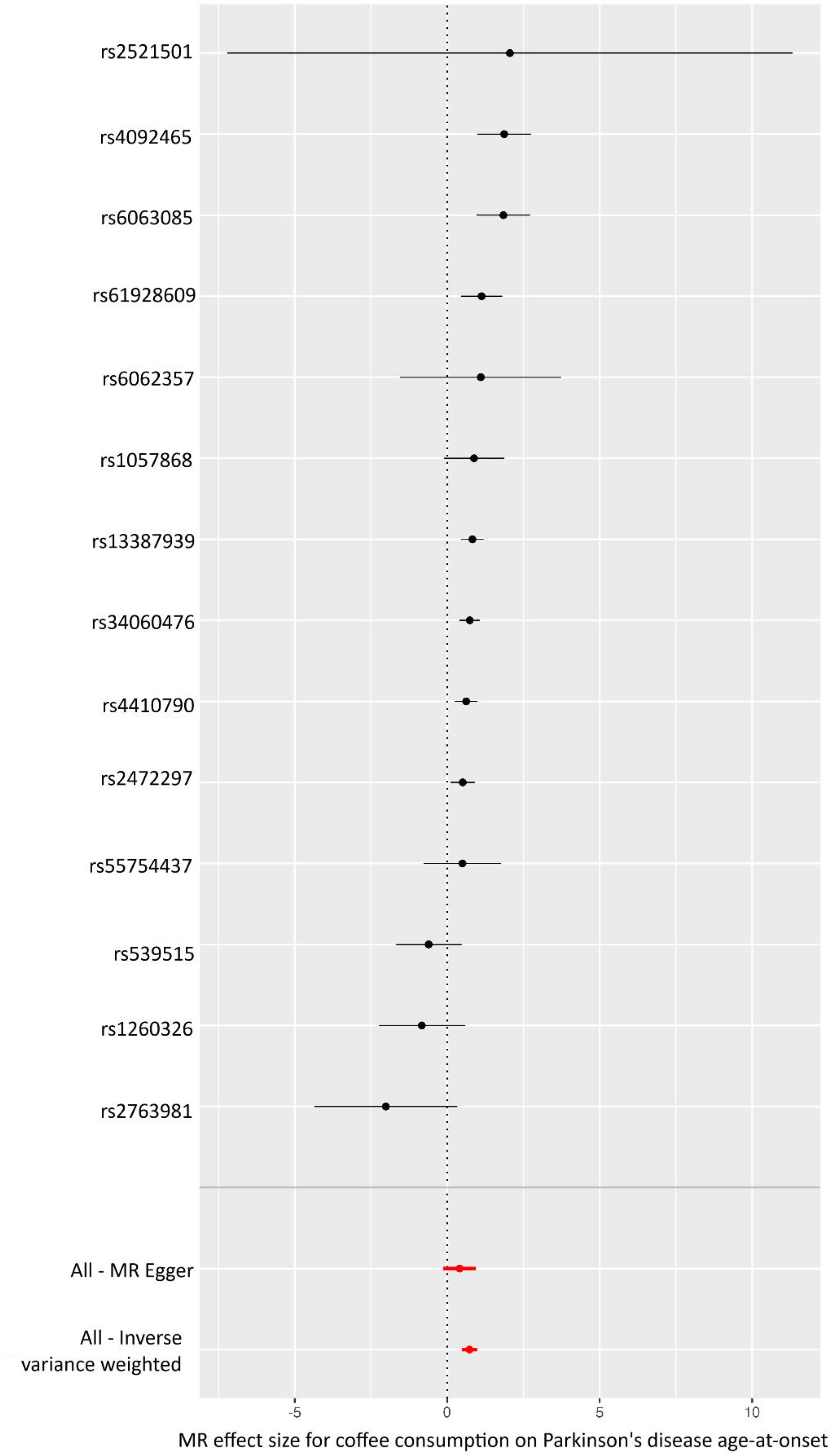
Forest plot of the MR analysis for the causal association between coffee consumption and the age at onset (AAO) of Parkinson’s disease (PD). Each point on the Y-axis corresponds to a single nucleotide polymorphism (SNP) used as an instrumental variable (IV). The X-axis shows the effect size (beta) for each SNP, with horizontal lines depicting 95% confidence intervals. In this scale, larger effect sizes indicate a later AAO (i.e., a delay in PD onset). The bottom two points display the overall MR-Egger and inverse variance– weighted (IVW) estimates, respectively, summarizing the combined effect of all IVs.

We then evaluated whether coffee consumption influences risk or clinical progression of PD, including motor (UPDRS3), cognitive (MMSE and MoCA), and non-motor features (hyposmia and sleep). Across all analyses, we observed no significant causal effects of coffee consumption **(Table 2)**. Sensitivity analyses further confirmed the robustness of these null results (**Supplementary Table 3**).

Finally, no significant genetic correlation between coffee consumption and PD risk or AAO was observed (**Supplementary Table 4**). PRS for coffee consumption also did not reveal significant associations with PD risk (OR = 1.02, 95% CI: 0.993–1.048, P = 0.147) or AAO (OR = 0.083, 95% CI: −0.108–0.274, P = 0.397; **Supplementary Table 5**).

## Discussion

Our findings indicate a potential protective relationship between coffee consumption and PD onset, suggesting that greater coffee intake may delay the emergence of clinical symptoms without altering overall disease risk or progression. After addressing pleiotropy in the MR analysis, we observed a robust association with later AAO, whereas neither MR, PRS nor genetic correlation analyses supported an effect on PD risk. These results align with previous reports linking coffee consumption to a delayed symptomatic onset of PD ^5, 17^ and further refine our understanding of coffee consumption role in PD, pointing toward mechanisms that modulate symptom onset rather than disease susceptibility.

One hypothesis is that the antagonistic effect of coffee on adenosine A2A receptors may help delay the emergence of overt motor symptoms. Notably, we identified several pleiotropic variants related to traits such as energy metabolism and smoking habits - one of SNPs was located near the *ADORA2A* receptor gene, yet none of these variants independently influence PD risk or AAO. Meanwhile, istradefylline, a selective adenosine A2A receptor antagonist, has been approved in several countries and shown efficacy in improving motor symptoms of PD ^18^. Earlier MR studies have likewise failed to establish a causal association between coffee intake and PD risk ^7, 8^, suggesting that the effect of coffee may indeed be selective for AAO.

Further research in prodromal cohorts, particularly those with REM sleep behavior disorder (RBD), is needed to clarify whether coffee consumption can delay or modify progression to PD. Although some evidence does not support a protective effect on RBD phenoconversion ^19, 20^, other findings suggest that RBD patients who subsequently developed parkinsonism rather than dementia consumed more caffeine ^19^. Meanwhile, one of the few studies conducted in genetically stratified cohorts demonstrated a protective effect of coffee in carriers of *LRRK2* variants ^21^, which is notable given that *LRRK2*-PD typically characterized by a lower prevalence of RBD and reduced dementia rates compared with sporadic or GBA-associated PD ^22^.

We acknowledge several limitations in our analysis. First, our study focused on individuals of European descent, which reflects the largest available GWAS data but limits the generalizability of our findings to other populations. Second, the available GWASs for PD progression phenotypes are relatively small and may be underpowered to detect an association. Third, even after excluding all detected pleiotropic SNPs from MR, residual pleiotropy cannot be completely ruled out. Furthermore, the protective signal may predominantly reflect specific genetic subtypes of PD, emphasizing the need to stratify PD by genetic subtypes in future research. Finally, we did not investigate potential sex-specific effects or interactions with specific therapies, which could influence disease onset and progression.

Overall, our findings suggest that coffee consumption appears to delay PD onset rather than reducing overall disease risk or modifying disease phenotype. Importantly, these results reflect the effect of genetically predicted coffee consumption on PD AAO as captured by MR and thus may not directly translate to increased coffee intake in everyday dietary practice. Future research should focus on validating these observations in broader and genetically diverse populations, ultimately aiming to clarify the mechanistic basis and therapeutic potential of coffee in PD.

## Data Availability

Summary statistics used in the analysis are publicly available. The cohorts used for PRS analysis are listed in Supplementary Table 1 and in Acknowledgements. Access to these cohorts is restricted to qualified researchers and can be requested through their respective data access portals or governing bodies.

## Acknowledgments

ZGO is supported by the Fonds de recherche du Québec - Santé (FRQS) Chercheurs-boursiers award, and is a William Dawson Scholar.

We would like to thank the participants in the different cohorts for contributing to this study. Data used in the preparation of this article were obtained from the Accelerating Medicine Partnership® (AMP®) Parkinson’s Disease (AMP PD) Knowledge Platform. For up-to-date information on the study, visit **https://www.amp-pd.org.** The AMP® PD program is a public-private partnership managed by the Foundation for the National Institutes of Health and funded by the National Institute of Neurological Disorders and Stroke (NINDS) in partnership with the Aligning Science Across Parkinson’s (ASAP) initiative; Celgene Corporation, a subsidiary of Bristol-Myers Squibb Company; GlaxoSmithKline plc (GSK); The Michael J. Fox Foundation for Parkinson’s Research; Pfizer Inc.; AbbVie Inc.; Sanofi US Services Inc.; and Verily Life Sciences. ACCELERATING MEDICINES PARTNERSHIP and AMP are registered service marks of the U.S. Department of Health and Human Services.

Clinical data and biosamples used in preparation of this article were obtained from the (i) Michael J. Fox Foundation for Parkinson’s Research (MJFF) and National Institutes of Neurological Disorders and Stroke (NINDS) BioFIND study, (ii) Harvard Biomarkers Study (HBS) and the Stephen & Denise Adams Center for Parkinson’s Disease Research of Yale School of Medicine (CPDR-Y), (iii) National Institute on Aging (NIA) International Lewy Body Dementia Genetics Consortium Genome Sequencing in Lewy Body Dementia Case-control Cohort (LBD), (iv) MJFF LRRK2 Cohort Consortium (LCC), (v) NINDS Parkinson’s Disease Biomarkers Program (PDBP), (vi) MJFF Parkinson’s Progression Markers Initiative (PPMI), and (vii) NINDS Study of Isradipine as a Disease-modifying Agent in Subjects With Early Parkinson Disease, Phase 3 (STEADY-PD3) and (viii) the NINDS Study of Urate Elevation in Parkinson’s Disease, Phase 3 (SURE-PD3).

BioFIND is sponsored by The Michael J. Fox Foundation for Parkinson’s Research (MJFF) with support from the National Institute for Neurological Disorders and Stroke (NINDS). The BioFIND Investigators have not participated in reviewing the data analysis or content of the manuscript. For up-to-date information on the study, visit michaeljfox.org/news/biofind. Genome sequence data for the Lewy body dementia case-control cohort were generated at the Intramural Research Program of the U.S. National Institutes of Health. The study was supported in part by the National Institute on Aging (program #: 1ZIAAG000935) and the National Institute of Neurological Disorders and Stroke (program #: 1ZIANS003154).

The Harvard Biomarker Study (HBS) is a collaboration of HBS investigators [full list of HBS investigators found at **https://www.bwhparkinsoncenter.org/biobank/** and funded through philanthropy and NIH and Non-NIH funding sources. The Stephen & Denise Adams Center for Parkinson’s Disease Research of Yale School of Medicine is funded through philanthropy and NIH and non-NIH funding sources. The HBS and CPDR-Y Investigators have not participated in reviewing the data analysis or content of the manuscript.

PPMI is sponsored by The Michael J. Fox Foundation for Parkinson’s Research and supported by a consortium of scientific partners: [list the full names of all of the PPMI funding partners found at **
https://www.ppmi-info.org/about-ppmi/who-we-are/study-sponsors**]. The PPMI investigators have not participated in reviewing the data analysis or content of the manuscript. For up-to-date information on the study, visit **www.ppmi-info.org**. The Parkinson’s Disease Biomarker Program (PDBP) consortium is supported by the National Institute of Neurological Disorders and Stroke (NINDS) at the National Institutes of Health. A full list of PDBP investigators can be found at **https://pdbp.ninds.nih.gov/policy**. The PDBP investigators have not participated in reviewing the data analysis or content of the manuscript. The Study of Isradipine as a Disease-modifying Agent in Subjects With Early Parkinson Disease, Phase 3 (STEADY-PD3) is funded by the National Institute of Neurological Disorders and Stroke (NINDS) at the National Institutes of Health with support from The Michael J. Fox Foundation and the Parkinson Study Group. For additional study information, visit **https://clinicaltrials.gov/ct2/show/study/NCT02168842**. The STEADY-PD3 investigators have not participated in reviewing the data analysis or content of the manuscript. The Study of Urate Elevation in Parkinson’s Disease, Phase 3 (SURE-PD3) is funded by the National Institute of Neurological Disorders and Stroke (NINDS) at the National Institutes of Health with support from The Michael J. Fox Foundation and the Parkinson Study Group. For additional study information, visit **https://clinicaltrials.gov/ct2/show/NCT02642393**. The SURE-PD3 investigators have not participated in reviewing the data analysis or content of the manuscript.

## Author Contributions

DK and KS contributed to the conception and design of the study; DK LL, ZGO and KS contributed to the acquisition, analysis of data and contributed to drafting the text and preparing the figures.

## Data availability

The code used for the analysis is available on GitHub: https://github.com/senkkon/coffee_PD/. Summary statistics used in the analysis are publicly available. The cohorts used for PRS analysis are listed in Supplementary Table 1 and in Acknowledgements. Access to these cohorts is restricted to qualified researchers and can be requested through their respective data access portals or governing bodies.

## Supporting information

**Supplementary Table 1**. Cohorts included in the polygenic risk score analysis of caffeine consumption.

**Supplementary Table 2**. Significant independent SNPs from coffee consumption GWAS (exposure)

**Supplementary Table 3**. Heterogeneity tests and tests for directional horizontal pleiotropy between coffee consumption and PD risk, AAO and progression

**Supplementary Table 4**. Genetic correlation between coffee consumption and PD risk and AAO.

**Supplementary Table 5**. PRS of caffeine consumption in PD risk and AAO

