## Supplementary Tables for "Coffee consumption is associated with later age-at-onset of Parkinson’s disease"

**Supplementary Table 1.** **Cohorts included in the polygenic risk score analysis of caffeine consumption.**

| Cohort | N total | Case | Control | Male, N (%) |
| --- | --- | --- | --- | --- |
| APDGC | 923 | 621 | 302 | 0.65 |
| McGill | 3651 | 2381 | 1270 | 0.41 |
| PPMI | 581 | 417 | 164 | 0.67 |
| IPDGC | 10799 | 5229 | 5480 | 0.59 |
| NGRC | 3940 | 1972 | 1968 | 0.53 |
| NIND | 1686 | 896 | 790 | 0.51 |
| AMP-PD | 5555 | 1963 | 3092 | 0.50 |

APDGC - Autopsy-Confirmed Parkinson Disease GWAS Consortium, McGill - McGill University Parkinson’s Cohort, PPMI - Parkinson's Progression Markers Initiative, IPDGC - International Parkinson Disease Genomics Consortium, NGRC - NeuroGenetics Research Consortium, NINDS - National Institute of Neurological Disorders and Stroke Repository Parkinson’s Disease Collection, AMP-PD - Accelerating Medicines Partnership Parkinson's disease

**Supplementary Table 2.** **Significant independent SNPs from Coffee consumption GWAS (exposure).**

| rsnumbers | MR with PD AAO* | Chr-position HG19 | Coffee consumption, P | PD AAO, P |
| --- | --- | --- | --- | --- |
| rs395815 | excluded | chr8:109156532 | 3.2e-09 | 0.67 |
| rs4410790 | included | chr7:17284577 | 1.9e-132 | 0.51 |
| rs2236955 | excluded | chr3:50429876 | 4.2e-08 | 0.45 |
| rs13387939 | included | chr2:637498 | 2.7e-13 | 0.79 |
| rs7224815 | excluded | chr17:17845800 | 5.7e-09 | 0.56 |
| rs4357572 | excluded | chr1:50576710 | 5.9e-11 | 0.91 |
| rs34060476 | included | chr7:73037956 | 1.2e-22 | 0.77 |
| rs2465054 | excluded | chr6:51174232 | 9.2e-09 | 0.48 |
| rs1422191 | excluded | chr5:87949158 | 4.1e-09 | 0.29 |
| rs55754437 | included | chr4:2933031 | 1.1e-08 | 0.90 |
| rs17004922 | excluded | chr22:24844948 | 4.7e-08 | 0.66 |
| rs6062357 | included | chr20:62892739 | 4.0e-11 | 0.79 |
| rs6063085 | excluded | chr20:45840459 | 4.8e-09 | 0.70 |
| rs13387939 | included | chr2:27730940 | 6.2e-17 | 0.77 |
| rs56113850 | excluded | chr19:41353107 | 3.2e-15 | 0.04 |
| rs66723169 | excluded | chr18:57808978 | 9.1e-19 | 0.35 |
| rs4092465 | excluded | chr18:55080437 | 3.3e-08 | 0.73 |
| rs62064918 | excluded | chr17:46155786 | 8.6e-09 | 0.08 |
| rs8056750 | excluded | chr16:70927078 | 1.5e-08 | 0.78 |
| rs17817964 | excluded | chr16:53828066 | 6.1e-17 | 0.28 |
| rs117968677 | excluded | chr15:75174251 | 5.1e-11 | 0.19 |
| rs2472297 | included | chr15:75027880 | 3.3e-168 | 0.57 |
| rs1057868 | included | chr7:75615006 | 7.6e-34 | 0.65 |
| rs2763981 | included | chr6:31840021 | 5.1e-10 | 0.68 |
| rs34190000 | excluded | chr5:7381260 | 6.5e-10 | 0.26 |
| rs2521501 | included | chr15:91437388 | 1.4e-08 | 0.63 |
| rs61928609 | included | chr12:11316437 | 1.2e-11 | 0.75 |
| rs539515 | included | chr1:177889025 | 2.7e-09 | 0.88 |

*Some SNPs were excluded from the Mendelian Randomization (MR) analysis on Parkinson’s disease (PD) age at onset (AAO) because they showed high pleiotropy as identified by MR-PRESSO.

**Supplementary Table 3.** **Heterogeneity tests and tests for directional horizontal pleiotropy between coffee consumption and PD risk, AAO and progression**

| Outcome | Heterogeneity tests | | | | | | Test for directional horizontal pleiotropy | | | |
| --- | --- | --- | --- | --- | --- | --- | --- | --- | --- | --- |
|  | MR Egger | | | Inverse variance weighted | | | egger_intercept | se | pval | MR-PRESSO global |
|  | Q | Q_df | Q_pval | Q | Q_df | Q_pval |  |  |  | pval |
| PD AAO | 18.289 | 10 | 0.051 | 18.35 | 11 | 0.074 | -0.003 | 0.017 | 0.851 | 0.202 |
| PD risk | 8.026 | 15 | 0.922 | 8.05 | 16 | 0.947 | 0.001 | 0.012 | 0.882 | 0.944 |
| UPDRS3 | 9.22 | 19 | 0.969 | 9.35 | 20 | 0.978 | 0.003 | 0.010 | 0.726 | 0.976 |
| MMSE | 0.236 | 2 | 0.889 | 0.25 | 3 | 0.970 | -0.005 | 0.051 | 0.928 | 0.976 |
| MoCa | 0.058 | 5 | 0.999 | 0.07 | 6 | 0.999 | 0.014 | 0.124 | 0.915 | 0.972 |
| Hyposomia | 9.224 | 19 | 0.970 | 9.35 | 20 | 0.979 | 0.004 | 0.010 | 0.726 | 0.974 |
| Sleep | 0.668 | 5 | 0.985 | 0.79 | 6 | 0.992 | -0.023 | 0.065 | 0.740 | 0.974 |

PD – Parkinson’s disease; AAO - Age at Onset; pval - p-value ; UPDRS3 - unified Parkinson's disease rating scale part 3; MMSE - Mini Mental State Examination; MoCA - Montreal Cognitive Assessment; OR - Odds ratio, L_CI95- low 95% confidence interval; U_CI95- upper 95% confidence interval

**Supplementary Table 4. Genetic correlation between coffee consumption and PD risk and AAO GWASs**

| **Outcome** | **rg** | **se** | **z** | **P** |
| --- | --- | --- | --- | --- |
| **PD risk** | 0.001 | 0.032 | 0.028 | 0.978 |
| **PD AAO** | -0.096 | 0.076 | -1.2592 | 0.208 |

PD – Parkinson’s disease; AAO – Age-at-onset; rg – genetic correlation coefficient; se – standard error.

**Supplementary Table 5. PRS of caffeine consumption in PD risk and AAO**

| PRS coffee consumption with PD risk | | | | |
| --- | --- | --- | --- | --- |
|  | OR | 95%-CI | %W(random) | p-value meta-analysis |
| Mcgill | 1.077 | [0.987;1.174] | 9.5 |  |
| PPMI | 1.026 | [0.845;1.247] | 1.9 |  |
| APDGC | 1.035 | [0.892;1.201] | 3.2 |  |
| IPDGC | 0.992 | [0.954;1.032] | 46.8 |  |
| NIND | 1.049 | [0.945;1.165] | 6.6 |  |
| NGRC | 1.048 | [0.974;1.128] | 13.4 |  |
| AMP PD | 1.023 | [0.968;1.096] | 18.5 |  |
| Meta-analysis Random effect model | 1.02 | [0.993;1.048] | | 0.147 |
| PRS coffee consumption with PD AAO | | | | |
|  | BETA | 95%-CI | %W(random) |  |
| Mcgill | -0.131 | [-0.556; 0.295] | 20.1 | p-value meta-analysis |
| PPMI | -0.086 | [-1.126; 0.953] | 3.4 |  |
| APDGC | 0.252 | [-0.423; 0.927] | 8 |  |
| IPDGC | -0.031 | [-0.378; 0.316] | 30.3 |  |
| NIND | 0.412 | [-0.328; 1.151] | 6.7 |  |
| NGRC | 0.368 | [-0.165; 0.901] | 12.8 |  |
| AMP PD | 0.141 | [-0.301; 0.583] | 18.6 |  |
| Meta-analysis Random effect model | 0.083 | [-0.1084; 0.2735] | | 0.397 |

OR – Odds Ratio, CI – Confidence Interval, PPMI – Parkinson's Progression Markers Initiative, APDGC – Autopsy-Confirmed Parkinson Disease GWAS Consortium, IPDGC – International Parkinson Disease Genomics Consortium, NINDS – National Institute of Neurological Disorders and Stroke Repository Parkinson's Disease Collection, NGRC – NeuroGenetics Research Consortium, AMP-PD - Accelerating Medicines Partnership Parkinson's disease
